# Post-Licensure Safety of Nirsevimab from the Canadian National Vaccine Safety (CANVAS) Network

**DOI:** 10.1101/2025.08.14.25333604

**Authors:** Marina Viñeta Paramo, Marilou Kiely, Louis Valiquette, Matthew P. Muller, Allison McGeer, Jennifer E. Isenor, Manish Sadarangani, James D. Kellner, Otto G. Vanderkooi, Kimberly Marty, Pascal M. Lavoie, Julie A. Bettinger, the Canadian Immunization Research Network

## Abstract

**Background and objectives:** Although clinical trials have shown that nirsevimab is safe, post-licensure safety data from routine settings are lacking. This study describes parent-reported health events occurring within seven days of nirsevimab administration in Canada.

**Methods:** Post-licensure, active, safety surveillance of nirsevimab during the 2024-25 viral season in Canada, based on parent-completed questionnaires seven days after immunization.

**Results:** Parents or caregivers of 1,559 children completed the survey. A third (454/1,559) of children had received nirsevimab co-administered with routine vaccines. Local injection-site reactions were reported in 140 children (9.0%), 79/454, 17.4%, [95%CI: 14.2–21.3] when nirsevimab was co-administered and 61/1,105, 5.5% [95%CI: 4.3–7.1] when nirsevimab was given alone. Injection-site reactions extending beyond the closest joint were infrequent (6/1,559, 0.4%). Health events that prevented daily activities or required a healthcare consultation were reported in 38/1,105 (3.4%) cases with nirsevimab administered alone and 19/454 (4.2%) with nirsevimab co-administered. The most reported symptoms were rhinorrhea: 1.8%, cough: 1.7%, feeding/eating changes: 1.6%, fever: 1.6% and diarrhea or change in bowel habits: 1.5%. Rash occurred in 10 children (0.6%). No cases of anaphylaxis were reported.

**Conclusions:** This study showed nirsevimab was well tolerated, with low incidence of health events within seven days of immunization. Results after co-administration support incorporation of nirsevimab into the routine vaccination schedule. Documenting parent-reported outcomes in post-licensure settings expands the safety data obtained from clinical trials and offers transparent, timely additional reassuring data to enhance parental confidence in RSV antibody interventions.

## INTRODUCTION

Respiratory syncytial virus (RSV) is the main cause of lower tract respiratory infections in young children, leading to hospitalization in 1 to 2% of children less than 2 years of age^1^. In 2023, the long-acting monoclonal antibody nirsevimab was approved and introduced by several jurisdictions for prevention of RSV disease in all infants and in high-risk children under 2 years of age^2–4^. Randomized controlled trials showed that nirsevimab administration is safe, with no significant difference in adverse events between treatment and placebo groups^5^. The majority of reported events, including local site reactions or fever, were mild or moderate, and were attributed to treatment in less than 2% of cases^5^. Despite these reassuring data, studies have shown that parental hesitancy towards this new intervention is mainly due to safety concerns, which can be exacerbated by a lack of independent, observational safety studies^6,7^. Currently, post-licensure safety surveillance studies are lacking to address these safety concerns.

The Canadian National Vaccine Safety (CANVAS) network (www.canvas-network.ca) is an active, participant-centered safety surveillance system for vaccines^8^, which also monitored the safety of nirsevimab for the 2024-2025 RSV season. Compared to other safety study designs, an active, participant-centered surveillance system offers several advantages: it typically includes a broader and more diverse population than clinical trials; it enables timely collection of acute events, thereby reducing recall bias compared to retrospective analysis; and it allows for the detection of a wider spectrum of adverse events, namely from mild to serious, transient, or even functional, including effects on daily activities, which may not be captured by healthcare practitioner reports. Moreover, by directly reflecting parent-perceived outcomes in routine practice rather than investigator-driven data, this approach provides independent information to understand vaccine tolerability, identify safety signals and inform family-centered care.

In Canada, RSV prevention programs with nirsevimab were initially deployed as universal infant programs in Quebec and Ontario in the fall of 2024, and as high-risk targeted program for other Canadian provinces^9–12^. The main objective of this study was to document parent-reported injection-site reactions and health events that interfered with daily activities or require medical consultation within seven days of nirsevimab administration. Secondary objectives were to examine rates of injection-site reactions and health events among children who received nirsevimab alone and those who received it alongside routine vaccines.

## METHODS

### Design and setting

This was a prospective, active surveillance study that enrolled children November 1, 2024 to March 31, 2025 and included children who received nirsevimab in the eastern provinces of Quebec, Ontario and Nova Scotia. In Quebec and Ontario, universal immunization was available for children born during the RSV season or less than 6 months old at the start of the season and children <24 months that remain at high-risk in their second season^9,10^. In Nova Scotia, nirsevimab was offered to children born ≤32 weeks’ gestation, with chronic lung disease or with hemodynamically significant congenital heart disease for their first RSV season and children with chronic lung disease for their second RSV season^11^.

### Study recruitment strategies

Different recruitment strategies were used among provinces^8^. In Quebec City and Sherbrooke (which are located in the province of Quebec), CANVAS sites automatically sent a study invitation to parents or caregivers of any child whose nirsevimab administration was entered into Quebec’s immunization registry and for whom the parent’s email address was available based on registry linkage. In Nova Scotia, parents or caregivers attending the RSV clinic for high-risk children in Halifax were invited to self-enroll by a research nurse. In the province of Ontario, passive recruitment used advertisements in hospital obstetric or pediatric clinics offering and administering nirsevimab. Parents or caregivers who consented to participate in the study received a link to the online safety questionnaire 8 days after immunization. Non-responders were sent a reminder survey link 3 and 6 days after the original link.

### Description of questionnaire

The online questionnaire collected information on child demographics (sex, age in months at time of immunization, race and ethnicity and administration of other vaccines concurrently with nirsevimab), on injection-site reactions and on new or worsening health problems that prevented daily activities or required healthcare consultation within seven days of nirsevimab administration (including a list of symptoms, onset and duration of symptoms, level of healthcare assistance sought and treatments received).

### Analytical outcomes

There were two primary outcomes for our analysis:

1. injection-related reactions: local (redness, pain or swelling at the injection site) and large (redness, pain or swelling above/below the closest joint in the immunized leg).
2. health events associated with new or worsening symptoms that occurred within 7 days after nirsevimab administration and were severe enough to prevent normal activities or to trigger a healthcare consultation (herein referred to as *health event*).

Secondary analytic outcomes were presence of *fever, rash or hives,* and *anaphylaxis* as typical symptoms for allergic reactions; and *serious health events*, defined as health events leading to an emergency department visit or to hospitalization.

### Statistical analysis

Baseline characteristics were summarized with median and interquartile range for continuous variables and with frequency and proportion for categorical variables. The proportions (%) of injection-related reactions and significant health events within 7 days of nirsevimab administration were calculated for the whole cohort and further stratified by children who received nirsevimab alone and those who received it concomitantly with routine vaccines. Ninety-five percent confidence intervals (95% CI) were estimated with the Wilson method^13^ for the primary and secondary outcomes.

To characterize the timing of health events following nirsevimab immunization, the onset and duration of events were summarized using ordinal time intervals and presented graphically. For individuals who reported “on-going” symptoms at day of questionnaire completion, the minimum duration was calculated as the days between onset and the day of completion. A survey was considered complete if the question “In the first week (7 days) after the dose of nirsevimab, did [first_name] develop a new health problem or did an existing health problem get worse?” was answered.

### Ethics statement

The Research Ethics Board at each participating site approved the study (Unity Health Toronto REB #20-334; Health Prince Edward Island and IWK Health Research REB #1026400; Centre Intégré univrsitaire de santé et de services sociaux de l’Estrie REB # F1H-63413).

## RESULTS

### Baseline characteristics

A total of 1,559 parents or caregivers completed the questionnaire, at a median time of 10 (IQR: 8-34) days after administration of nirsevimab. In Quebec and Nova Scotia, active recruitment yielded a response rate of 26.3% (1,546 of 5,871 invitations) and 28.6% (4 of 14 invitations), respectively. Since Ontario used passive recruitment, the response rate could not be ascertained. Respondent rates and number of doses given per province can be found in Supplemental table 1. Most children received nirsevimab before reaching 7 months of age. The sample had an even sex distribution, with most respondents residing in Quebec. Most parents or caregivers completed the survey in French and identified their child as being of White origin (Table 1). Nirsevimab was co-administered with routine vaccines in just under a third (454 of 1,559; 29.1%) of children. Details on the specific vaccines co-administered are provided in Supplemental table 2.

**Table 1.**
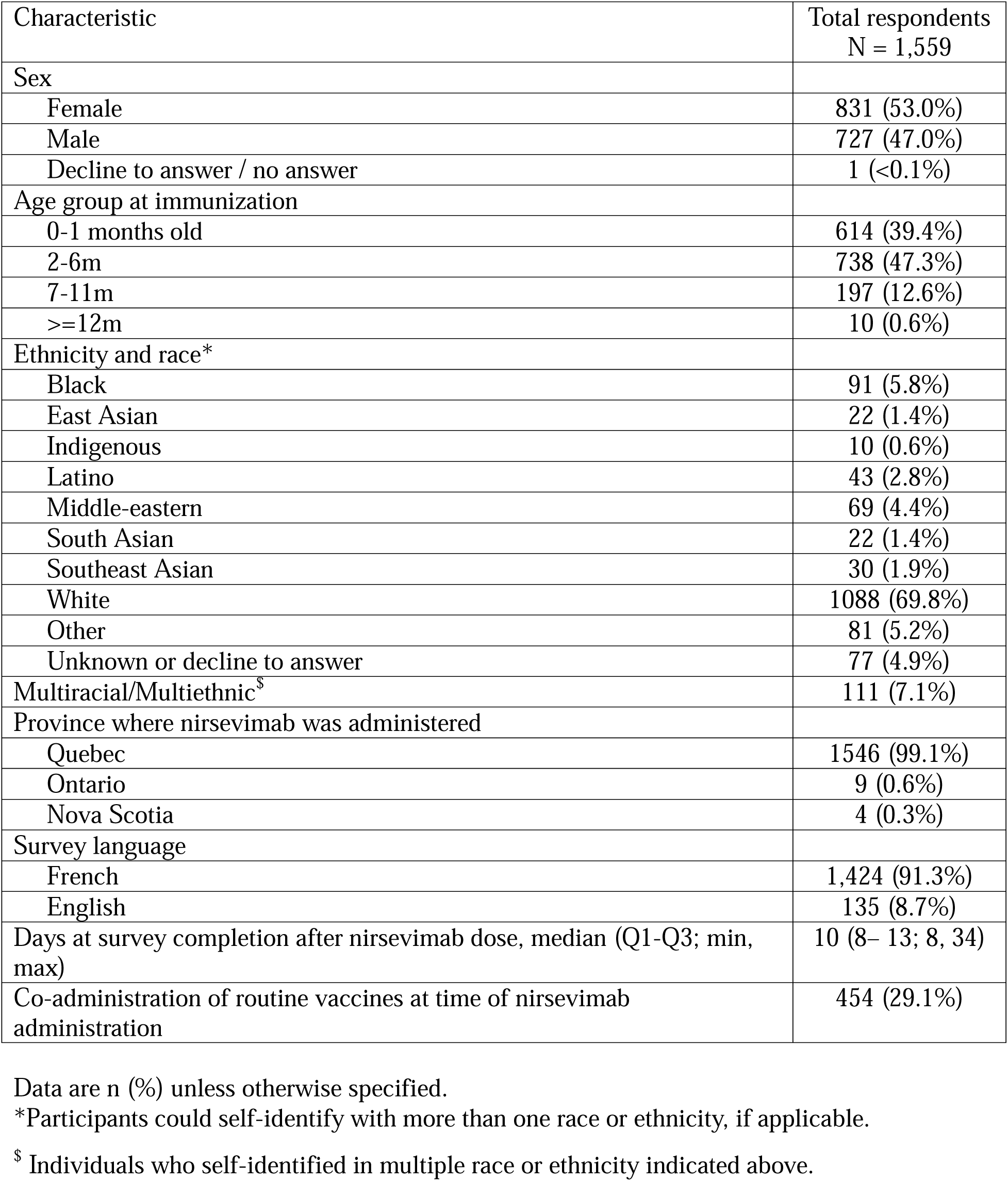
Baseline demographics of the study population.

| Characteristic | Total respondents<br>N = 1,559 |
| --- | --- |
| **Sex** |  |
| Female | 831 (53.0%) |
| Male | 727 (47.0%) |
| Decline to answer / no answer | 1 (<0.1%) |
| **Age group at immunization** |  |
| 0-1 months old | 614 (39.4%) |
| 2-6m | 738 (47.3%) |
| 7-11m | 197 (12.6%) |
| >=12m | 10 (0.6%) |
| **Ethnicity and race\*** |  |
| Black | 91 (5.8%) |
| East Asian | 22 (1.4%) |
| Indigenous | 10 (0.6%) |
| Latino | 43 (2.8%) |
| Middle-eastern | 69 (4.4%) |
| South Asian | 22 (1.4%) |
| Southeast Asian | 30 (1.9%) |
| White | 1088 (69.8%) |
| Other | 81 (5.2%) |
| Unknown or decline to answer | 77 (4.9%) |
| Multiracial/Multiethnic<sup>\$</sup> | 111 (7.1%) |
| **Province where nirsevimab was administered** |  |
| Quebec | 1546 (99.1%) |
| Ontario | 9 (0.6%) |
| Nova Scotia | 4 (0.3%) |
| **Survey language** |  |
| French | 1,424 (91.3%) |
| English | 135 (8.7%) |
| Days at survey completion after nirsevimab dose, median (Q1-Q3; min, max) | 10 (8– 13; 8, 34) |
| Co-administration of routine vaccines at time of nirsevimab administration | 454 (29.1%) |

Data are n (%) unless otherwise specified.
\*Participants could self-identify with more than one race or ethnicity, if applicable.
<sup>\$</sup> Individuals who self-identified in multiple race or ethnicity indicated above.

### Main outcomes

Local injection-site reactions were reported in 140 (9.0%) children, specifically in 5.5% (95% CI: 4.3 – 7.1) of the children who received nirsevimab alone and 17.4% (95% CI: 14.2 – 21.3) of those who received it alongside other routine vaccines (Table 2). Large injection-site reactions extending beyond the nearest joint were infrequent in children who received nirsevimab alone and those who received it with routine vaccines (Table 2).

**Table 2.**
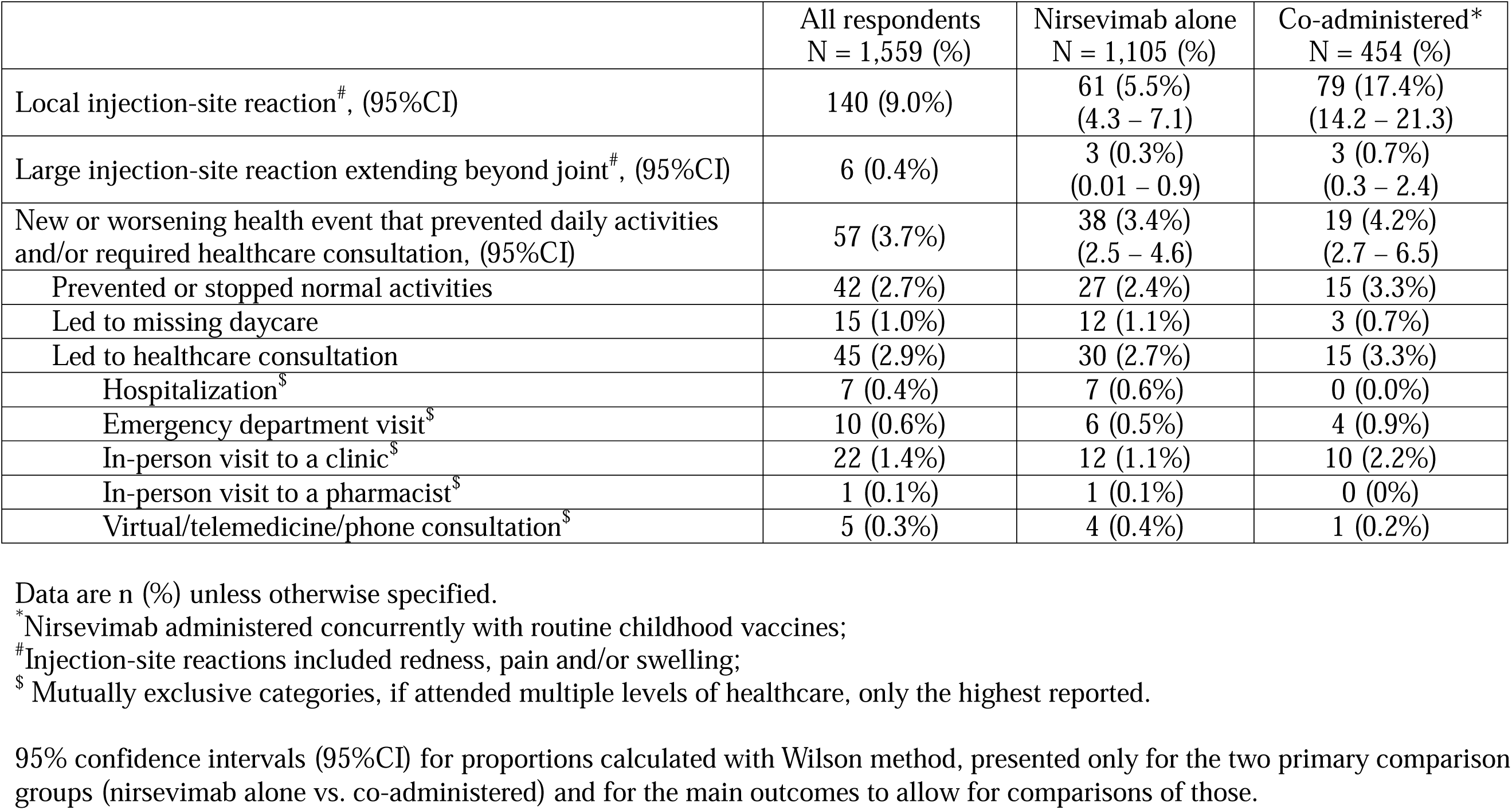
Injection-site reactions and health events within 7 days of nirsevimab administration, alone or co-administered with routine childhood vaccines.

|  | All respondents<br>N = 1,559 (%) | Nirsevimab alone<br>N = 1,105 (%) | Co-administered*<br>N = 454 (%) |
| --- | --- | --- | --- |
| Local injection-site reaction<sup>#</sup>, (95%CI) | 140 (9.0%) | 61 (5.5%)<br>(4.3 – 7.1) | 79 (17.4%)<br>(14.2 – 21.3) |
| Large injection-site reaction extending beyond joint<sup>#</sup>, (95%CI) | 6 (0.4%) | 3 (0.3%)<br>(0.01 – 0.9) | 3 (0.7%)<br>(0.3 – 2.4) |
| New or worsening health event that prevented daily activities and/or required healthcare consultation, (95%CI) | 57 (3.7%) | 38 (3.4%)<br>(2.5 – 4.6) | 19 (4.2%)<br>(2.7 – 6.5) |
| Prevented or stopped normal activities | 42 (2.7%) | 27 (2.4%) | 15 (3.3%) |
| Led to missing daycare | 15 (1.0%) | 12 (1.1%) | 3 (0.7%) |
| Led to healthcare consultation | 45 (2.9%) | 30 (2.7%) | 15 (3.3%) |
| Hospitalization<sup>\$</sup> | 7 (0.4%) | 7 (0.6%) | 0 (0.0%) |
| Emergency department visit<sup>\$</sup> | 10 (0.6%) | 6 (0.5%) | 4 (0.9%) |
| In-person visit to a clinic<sup>\$</sup> | 22 (1.4%) | 12 (1.1%) | 10 (2.2%) |
| In-person visit to a pharmacist<sup>\$</sup> | 1 (0.1%) | 1 (0.1%) | 0 (0%) |
| Virtual/telemedicine/phone consultation<sup>\$</sup> | 5 (0.3%) | 4 (0.4%) | 1 (0.2%) |

Data are n (%) unless otherwise specified.
\*Nirsevimab administered concurrently with routine childhood vaccines;
<sup>#</sup>Injection-site reactions included redness, pain and/or swelling;
<sup>\$</sup> Mutually exclusive categories, if attended multiple levels of healthcare, only the highest reported.
95% confidence intervals (95%CI) for proportions calculated with Wilson method, presented only for the two primary comparison groups (nirsevimab alone vs. co-administered) and for the main outcomes to allow for comparisons of those.

A new or worsening health event that prevented daily activities and/or required healthcare consultation was reported in 57 (3.7%) of respondents, with corresponding rates of 3.4% and 4.2% in children who received nirsevimab alone or with routine vaccines, respectively (Table 2).

### Symptoms and timing of health events occurring after immunization

Rhinorrhea, cough, nasal or sinus congestion, changes in eating/feeding pattern, and fever were the most frequently reported symptoms, occurring in more than 1.5% of respondents (Table 3). The proportion of children with cough, nasal congestion, diarrhea and vomiting was approximately twice as high among those who received nirsevimab alongside routine vaccines compared to those who received it alone (Table 3). Fever was reported in 1.4% (95% CI: 0.9 – 2.3) of children who received nirsevimab alone and in 2.0% (95% CI: 1.1 – 3.8) of those who received it with routine vaccines. Rash or hives occurred in 0.5% (95% CI: 0.2 – 1.1) of children in the nirsevimab-alone group and 0.9% (95% CI: 0.4 – 2.3) in the co-administration group (Table 3). Although anaphylaxis was specifically solicited, no cases were reported.

**Table 3.**
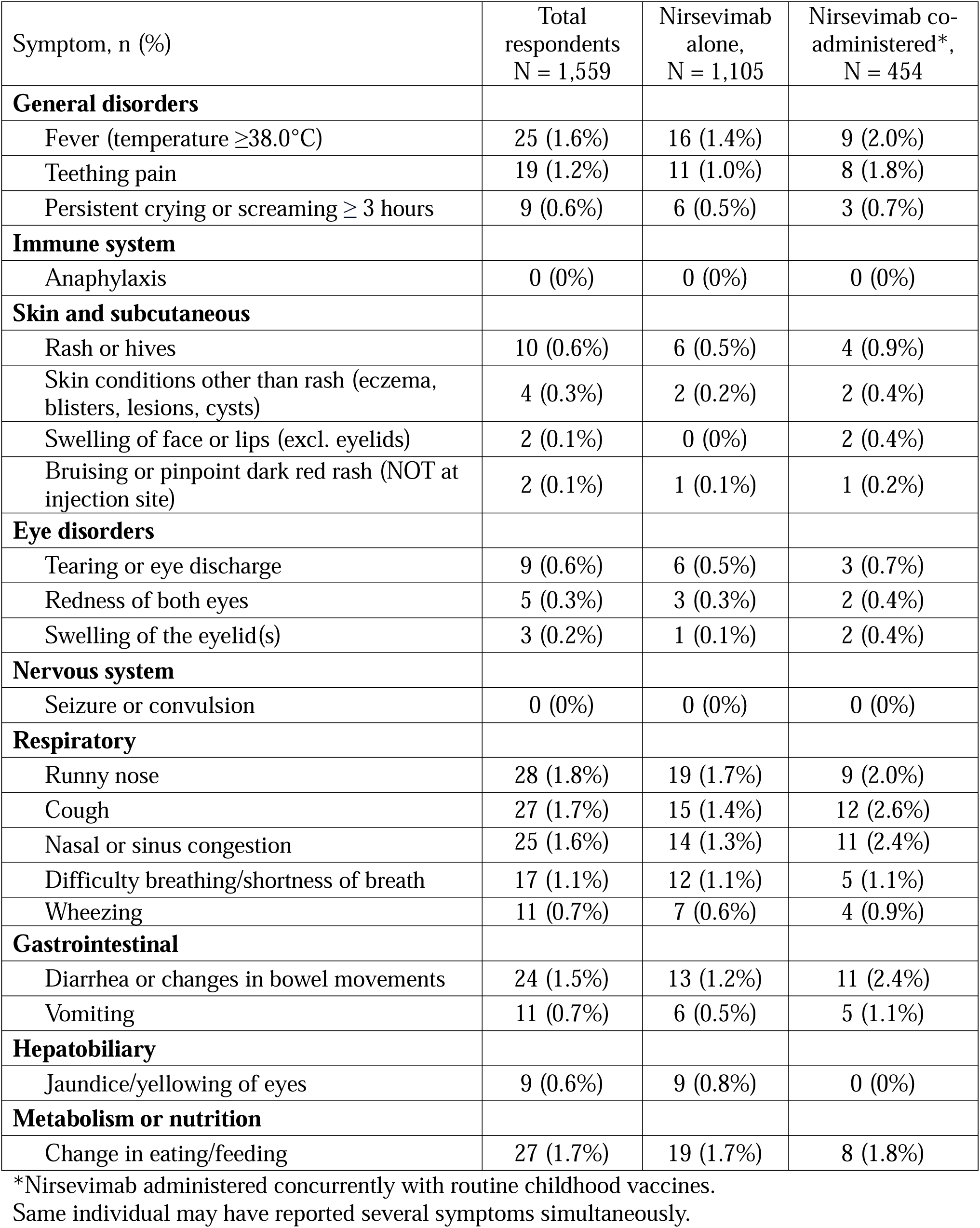
Symptoms associated with health events within 7 days of nirsevimab administration, alone or co-administered with routine childhood vaccines.

| Symptom, n (%) | Total respondents<br>N = 1,559 | Nirsevimab alone,<br>N = 1,105 | Nirsevimab co-administered*,<br>N = 454 |
| --- | --- | --- | --- |
| **General disorders** |  |  |  |
| Fever (temperature $\geq 38.0^{\circ}\text{C}$) | 25 (1.6%) | 16 (1.4%) | 9 (2.0%) |
| Teething pain | 19 (1.2%) | 11 (1.0%) | 8 (1.8%) |
| Persistent crying or screaming $\geq 3$ hours | 9 (0.6%) | 6 (0.5%) | 3 (0.7%) |
| **Immune system** |  |  |  |
| Anaphylaxis | 0 (0%) | 0 (0%) | 0 (0%) |
| **Skin and subcutaneous** |  |  |  |
| Rash or hives | 10 (0.6%) | 6 (0.5%) | 4 (0.9%) |
| Skin conditions other than rash (eczema, blisters, lesions, cysts) | 4 (0.3%) | 2 (0.2%) | 2 (0.4%) |
| Swelling of face or lips (excl. eyelids) | 2 (0.1%) | 0 (0%) | 2 (0.4%) |
| Bruising or pinpoint dark red rash (NOT at injection site) | 2 (0.1%) | 1 (0.1%) | 1 (0.2%) |
| **Eye disorders** |  |  |  |
| Tearing or eye discharge | 9 (0.6%) | 6 (0.5%) | 3 (0.7%) |
| Redness of both eyes | 5 (0.3%) | 3 (0.3%) | 2 (0.4%) |
| Swelling of the eyelid(s) | 3 (0.2%) | 1 (0.1%) | 2 (0.4%) |
| **Nervous system** |  |  |  |
| Seizure or convulsion | 0 (0%) | 0 (0%) | 0 (0%) |
| **Respiratory** |  |  |  |
| Runny nose | 28 (1.8%) | 19 (1.7%) | 9 (2.0%) |
| Cough | 27 (1.7%) | 15 (1.4%) | 12 (2.6%) |
| Nasal or sinus congestion | 25 (1.6%) | 14 (1.3%) | 11 (2.4%) |
| Difficulty breathing/shortness of breath | 17 (1.1%) | 12 (1.1%) | 5 (1.1%) |
| Wheezing | 11 (0.7%) | 7 (0.6%) | 4 (0.9%) |
| **Gastrointestinal** |  |  |  |
| Diarrhea or changes in bowel movements | 24 (1.5%) | 13 (1.2%) | 11 (2.4%) |
| Vomiting | 11 (0.7%) | 6 (0.5%) | 5 (1.1%) |
| **Hepatobiliary** |  |  |  |
| Jaundice/yellowing of eyes | 9 (0.6%) | 9 (0.8%) | 0 (0%) |
| **Metabolism or nutrition** |  |  |  |
| Change in eating/feeding | 27 (1.7%) | 19 (1.7%) | 8 (1.8%) |

\*Nirsevimab administered concurrently with routine childhood vaccines.
Same individual may have reported several symptoms simultaneously.

Most health events began 2 to 3 days after nirsevimab administration (26 of 57 health events; 45.6%), with same distribution between children who receiving nirsevimab alone and those who received it with routine vaccines (Figure 1A). Among 16 health events with onset within 24 hours of immunization, the most concerning reported symptom was respiratory issues (n=5), gastrointestinal symptoms (n=6), persistent crying or change in feeding (n=2), rash or hives (n=1), face swelling (n=1) and hypothermia (n=1).

**Figure 1.**
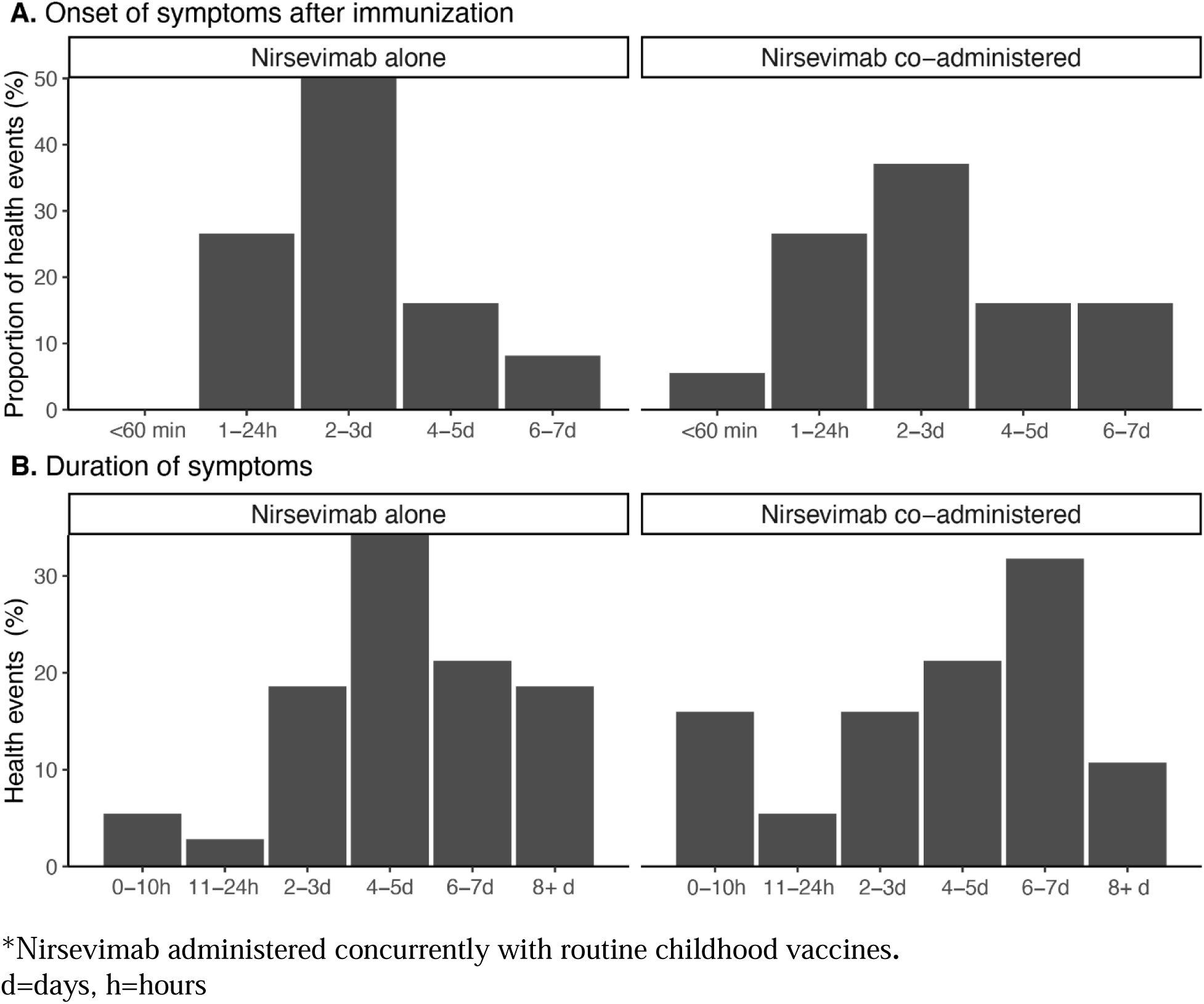
Onset (A, B) and duration (C, D) of health events within 7 days of nirsevimab administration, alone or co-administered with routine childhood vaccines.

Among events that lasted 4 days or more (n=40) the most concerning symptom reported was respiratory (19 of 40 events, 47.5%); whereas among 17 events that lasted up to 3 days, the most concerning symptom reported was fever (5 of 17 events, 29.4%), followed by gastrointestinal symptoms (3 of 17 events, 17.6%).

### Serious health events, requiring emergency department visit or hospitalization

Ten children (0.6% of respondents) attended an emergency department. Among those children, four reported diagnosed or presumptive acute viral infections (RSV infection, pneumonia, laryngitis and conjunctivitis). Five children did not report a specific medical diagnosis, but reported the following as their most concerning symptom (n=1 each): diarrhea, difficulty breathing, fever, cough and persistent crying.

The remaining child who visited the emergency department received a medical diagnosis of “post-vaccination allergic reaction”, but did not require specific treatment. This was a 2-month-old female who had received nirsevimab concurrently with the 2-month routine vaccines (Infanrix hexa^®^ [diphtheria, tetanus, acellular pertussis, hepatitis B, inactivated poliovirus and *Haemophilus influenza*e type B], Vaxneuvance^®^ [15-valent pneumococcal conjugate vaccine] and RotaTeq^®^ [Rotavirus pentavalent vaccine]). Onset of symptoms was 60 minutes after immunization/vaccination and symptoms lasted for 10 hours. The main symptom reported was facial swelling, associated with rash, eye redness and swelling, nasal congestion, runny nose and coughing. A skin reaction, without respiratory symptoms or facial swelling occurred 24 hours after the 4-month vaccine series administration. The case was followed by an allergist, who did not diagnose a systemic vaccine allergy, and by provincial public health, who recommended subsequent vaccines be administered according to schedule under medical supervision.

Six hospital admissions occurred in infants younger than 1 month of age and one in a 2-month-old infant, all after sole administration of nirsevimab. Three hospitalized infants had a diagnosis of neonatal jaundice; two had respiratory viral illnesses (RSV and influenza); one neonate was hospitalized for hypothermia; and one child did not receive a medical diagnosis but reported nasal congestion as main symptom, combined with difficulty breathing and wheezing, without any dermatological or gastrointestinal symptoms, that would indicate an allergic reaction. None required intensive care unit admission. Five cases had resolved and been discharged home and one was still hospitalized at survey completion.

## DISCUSSION

In this active surveillance study, we described the safety profile of nirsevimab in Canadian infants, as reported by parents and caregivers. The most common event was local injection-site reactions which occurred more frequently in children who received nirsevimab concurrently with other routine childhood vaccines. Health events that prevented daily activities or required healthcare consultation in the 7 days after nirsevimab administration were infrequent (3.7%), with no apparent differences between children who received nirsevimab alone versus those who received it concurrently with routine vaccines. Overall, the data depict a favorable tolerability profile for nirsevimab, with mild and temporary health events following immunization.

A key strength of this study is the reporting of outcomes through the CANVAS network, a participant-centered, active safety surveillance platform that enables timely capture of parent-reported health events. Unlike passive surveillance systems that primarily capture medically attended adverse events, this study systematically collected information on a wide range of health outcomes of direct relevance to families. The CANVAS system can also detect safety signals that may be further explored with other study designs. Another strength is most participants were recruited via auto-enrollment from centralized vaccine registries. As shown previously, this allows for improved data quality on product administration information (e.g., date received, lot and type of product)^8^. It also ensures broader representativeness of the studied population across immunized children, since it removes barriers to participation by increasing awareness among all eligible individuals, achieving a larger and more balanced sample.^8^ Based on the limited number of previously published post-licensure safety studies, injection-site reactions with nirsevimab are anticipated to occur in 4% to 10% of children^14,15^, which is consistent with the 9.0% observed rate in this study. This contrast with the clinical trial data, particularly the pooled safety summary from the phase 2 and 3 nirsevimab trials which reported a much lower rate of 0.3% for injection-site reactions^5^. Interestingly, this rate matches the frequency of large injection-site reactions observed in our study, suggesting that trial data may capture only reactogenic events considered more significant by study staff, whereas our findings may more closely reflect the parent’s and caregiver’s perception of these health events.

This study adds new evidence on the safety of co-administering nirsevimab with routine childhood vaccines. The frequency of local reactions observed in this study is consistent with active safety surveillance data from Western Australia.^15^ They found that 2.3% of children receiving nirsevimab alone and 16.7% of those who received it with one or more routine vaccines experienced local site reactions. Compared to routine childhood vaccination at 2 and 4 months, adding nirsevimab does not increase the expected rate of local site reactions, which are estimated to be 10% to 16% at each time point, respectively, based on active surveillance^16,17^. Our study strengthens and enhances the generalization of data from these previous studies.

Fever, change in feeding, respiratory and gastrointestinal symptoms were the most frequently reported symptoms after nirsevimab immunization. A pooled analysis of nirsevimab RCTs, which included unimmunized controls found that the most frequent treatment-related events within seven days of injection were pyrexia and vomiting, with similar rates in the nirsevimab and placebo recipients^5^. The active surveillance study in Western Australia likewise identified fever and gastrointestinal symptoms as the most frequent, along with fatigue. Both our study and the Australian surveillance found that these events occurred more often when nirsevimab was co-administered with other vaccines. However, rates with co-administration did not exceed those expected for routine childhood vaccines at 2 and 4 months of age, were fever occurs in up to 16% of infants and gastrointestinal symptoms in up to 13%^16,17^. Altogether, this supports the feasibility of integrated immunization strategies and may help address concerns about the cumulative burden of infant vaccination if nirsevimab is incorporated into the routine schedule.

Serious health events, including emergency department visits and hospitalizations were rare and appeared mostly unrelated to immunization. Similar to other studies,^5,18,19^ no cases of anaphylaxis were reported. However, in our study one child was diagnosed with a “post-vaccination allergic reaction” after receiving nirsevimab concurrently with other routine vaccines. Since anaphylaxis and other hypersensitivity reactions are very rare (occurring in less than 0.01% of cases), the sample size of this study remains insufficient to estimate these risks, warranting continued monitoring of allergic reactions on a larger scale.

This study provides important information to support family discussion around immunization decisions. In a Spanish post-nirsevimab administration survey that included 1,692 respondents, 14 (0.8 %) were not satisfied with the immunization and about 1.5% would not recommend it or administer it to another child, mainly “due to side effects.”^20^ Similarly, a study in France highlighted that among parents who declined nirsevimab, 35% were concerned about the absence of long-term safety data, 20% objected to the intramuscular route, and 15% feared side effects.^6^ These concerns persist despite the availability of drug agency-approved monographs and detailed clinical-trial safety summaries.^5,21^ Two studies on childhood vaccine communication highlighted the importance of knowledge translation by (1) incorporating relatable parental narratives and (2) tailoring content to the specific concerns of each community.^22,23^ This underscores the need to provide transparent data, translating scientific evidence into accessible, and family-centred messages. The data presented here offer independent safety data at arm’s length from industry-funded trials, focused on the perspective of parents and caregivers, thus providing what families may consider a balanced source of information for family counselling on nirsevimab safety.^24^

This study has some limitations. First, it focuses on early-onset health events post-immunization; longitudinal cohort studies based on administrative datasets would be more suitable for the assessment of serious late onset events^25^. Second, the absence of a comparator group precludes causal analyses. Additionally, without a control group we cannot fully exclude a respondent bias towards those who experienced a health event, which would lead to overestimation of true rates in our study. Third, the study sample size was not powered to detect events occurring in less 1 in 400, impeding the assessment of uncommon events (event occurring in 0.1% to less than 1%)^26^. Finally, the study sample lacked demographic diversity. Most participants were recruited in Quebec and approximately two-thirds reported being White. While this race distribution is representative of the overall Canadian context^27^, and Quebec, future studies should intentionally expand recruitment in under-represented racial/ethnic groups to enhance external validity across Canada’s diverse population.

## CONCLUSION

This participant-centered active surveillance study found that nirsevimab was well tolerated among Canadian infants, with a low incidence of parent-reported health events within seven days of immunization. The rate of local injection-site reactions after sole administration was lower than that reported in clinical trials and lower than rates observed with routine infant vaccines. When co-administered with routine vaccines, the rate was comparable to that of routine vaccines alone, supporting the feasibility of co-administration. These findings extend the safety profile of nirsevimab beyond clinical trials and provide real-world evidence supporting its use in the general population, including high-risk children. By capturing parent-reported acute outcomes, this study helps fill gaps in post-licensure safety data, support generalizability of safety events reported in the limited number of active surveillance studies reported today and offers valuable insights to guide clinical practice and vaccine communication strategies.

## Data Availability

Deidentified individual participant data will not be made available because we do not have permission from CANVAS participants to share the data used in our study

## Abbreviations

(RSV): Respiratory syncytial virus
(CANVAS): Canadian National Vaccine Safety

## Acknowledgements

We would like to express our sincere appreciation to all study participants who made this study possible. Thanks to our provincial/territorial collaborators and other collaborators from Canadian Immunization Research Network.

## Contributors’ statement

JAB conceptualized and designed the study, supervised the development of analysis plan and critically reviewed and revised the manuscript for important intellectual content.

PML supervised the development of analysis plan, drafted the manuscript and critically reviewed and revised the manuscript for important intellectual content.

MVP developed the analysis plan and data cleaning analyzed, produced data tables and figures, drafted the initial manuscript and critically reviewed and revised the manuscript.

MPM, JDK, JEI, MK, KM, JDK, MS, AM, OGV, and LV contributed to conceptualization and design of the study and study instruments, coordinated and supervised study implementation and data collection, and critically reviewed and revised the manuscript for important intellectual content.

All authors approved the final manuscript as submitted and agree to be accountable for all aspects of the work.

## SUPPLEMENTAL MATERIAL

**Supplemental table 1:** Response rates and number of nirsevimab doses administered per province.

|  | Doses administered<sup>#</sup> | Families invited to participate* | Respondents, n (% of invitations) |
| --- | --- | --- | --- |
| Nova Scotia | 20 | 14 | 4 (28.6) |
| Ontario | NA | NA | 9 (NA) |
| Quebec | 30,951 | 5,871 | 1,546 (26.3) |

<sup>#</sup> Doses administered in recruitment site catchment area during study period
\*Available in Quebec where recruitment occurred via auto-enrollment based on e-mail availability in administrative datasets and in Nova Scotia, where active recruitment was conducted by a research nurse.
NA: Not available

**Supplemental table 2.** Type of vaccines co-administered at time of nirsevimab administration.

|  | Total respondents<br>N = 1559 |
| --- | --- |
| Co-administration of other vaccines at time of nirsevimab administration | 454 (29.1%) |
| 2-month routine vaccines<sup>1</sup> | 136 |
| 4-month routine vaccines<sup>1</sup> | 276* |
| 4CMenB | 4 |
| Influenza | 21 |
| COVID-19 | 2 |
| COVID-19 and influenza | 6 |
| Measles, mumps and rubella | 4 |
| No information on other vaccines administered | 6 |

<sup>1</sup> Include: Infanrix hexa<sup>®</sup> [diphtheria, tetanus, acellular pertussis, hepatitis B, inactivated poliovirus and *Haemophilus influenzae* type B], Vaxneuvance<sup>®</sup> [15-valent pneumococcal conjugate vaccine] and RotaTeq<sup>®</sup> [Rotavirus pentavalent vaccine]
\* One child received the pentavalent DTaP-Polio-Hib vaccine instead of Infanrix hexa

